# Management of Moderate-to-highly Exuding Leg Ulcers with Superabsorbent Wound Dressings versus Foams Dressings in Spanish Settings: An Early-stage Cost-effectiveness and Budget-impact Analyses

**DOI:** 10.1101/2023.05.19.23290229

**Authors:** Vladica Veličković, Joan-Enric Torra i Bou, Francisco Cegri, Federico Palomar Llatas

**Author notes:** **Author Approval** Please indicate that all authors have seen and approved the manuscript. **Data Availability Statement:** Data not available.

## Abstract

This study endeavors to ascertain the cost-effectiveness and utility of employing SAPs in comparison to foam dressings for the management of moderate-to-highly exuding leg ulcers within Spanish healthcare contexts. Furthermore, it aims to conduct an associated budget-impact analysis for the Spanish National Health Service, thereby considering the financial implications relevant to the implementation of these interventions.

The economic evaluation and budget impact model adhered to Spanish and international guidelines. with reporting quality of economic evaluation assessed using the CHEERS checklist and the Drummond checklist for ensuring adequate conduct. The evidence synthesis process involved identifying and appraising the most recent and highest level of evidence research. The identified data inputs were adjusted in accordance with recommended practices for research quality and incorporated into a mathematical model that accounted for the natural history of chronic leg ulcers.

Based on the results of this preliminary health economic analysis, utilizing SAP wound dressings instead of foam dressings in patients with moderate-to-highly exuding leg ulcers within the Spanish Health Care System is projected to yield favorable outcomes. The analysis predicts an enhanced healing rate of 2.33%, an incremental improvement in health-related quality of life (HRQoL) by 0.129 quality-adjusted life weeks (QALWs), and a total direct cost reduction of €570 per patient over a six-month period. Extrapolating these findings to a scenario where all leg ulcers are treated with SAPs rather than foam dressings, the Spanish NHS could potentially achieve an annual cost savings of €43.46 million.

These clinical outcomes align with current treatment guidelines that advocate for superabsorbent wound dressings as the primary choice for managing moderate-to-highly exuding leg ulcers.

## Introduction

Leg ulcers (LUs) represent a significant medical burden as they are the most frequent type of hard-to-heal ulcers. The etiology of LUs is mainly attributed to venous hypertension, venous insufficiency, obesity, and deep vein thrombosis (1). These ulcers tend to occur more frequently in the medial side of the lower leg, specifically between the ankle and the knee, and they exhibit little progress towards healing within a period of 4-6 weeks from the onset, leading to a chronic state (2). The incidence of LUs is higher among individuals aged 65 and older, which has become a concerning issue in light of the growing aging population (3). Diaz-Herrera et al. (4) examines the prevalence of chronic wounds in primary health in the Southern Metropolitan Area of Barcelona and he concludes that the more observed wounds in primary care setting are LUs (0.04%) compared with diabetic foot ulcers (0.015%) and pressure ulcers (0,033%). Berenguer et al. (5) estimated 0.8 (2010), 1.1 (2011), 1.6 (2012), 2.2 (2013), 2.1 (2014) cases per 1000 people per year respectively for 2010-2014. from real-world data in Spain. According to Verdú, J. et al. (6) LUs general population prevalence is from 0.8 to 0.5%, incidence between 2 and 5 new cases per thousand people per year. In addition, the average age patients is 52.3 +/- 18.5 in women and 51.9 +/- 18.9 in men 3% of patients with CVI (chronic venous insufficiency) have a venous ulcer, and 1% of patients with CVI have a grade 6 classification (CEAP system) (7). Regarding economic burden of chronic wounds, not many literatures on cost-of-illness studies has been found in Spain. Agreda et al. (8) estimates the economic impact of pressure ulcer management, but overall cost of leg ulcers remains unknown. On the other hand, in Germany, the estimated cost per patient for treating a LU is €6.905 (9), whereas studies in the UK suggest mean costs of £5.488 per patient (10).

Chronic wounds are treated according to the TIMERS principle (11), which provides structured guidance on wound management and identifies where advanced adjunctive therapies may be used. The framework considers tissue viability, infection/inflammation, moisture balance, wound edge, repair/regeneration, and social- and patient-related factors. Dressings are the mainstay of wound exudate management, the aim of which is to optimise wound bed moisture levels, protect the surrounding skin, and manage symptoms and improve patient Quality of Life (QoL) (12). Dressing selection should consider local factors, e.g., infection, which may require a dressing which can deliver topical antimicrobials. Dressings commonly used for the management of moderate-to-high exuding wounds are foams, carboxymethylcellulose (CMC), alginates, and super absorbent polymers (SAPs) (12). Primary dressings are those that are in direct contact with the wound and may require a separate method of fixation (12). Secondary dressings can be applied over the top of primary dressings to provide additional benefit. When choosing a dressing specifically for moderate to highly exuding LUs, various dressing properties should be considered. Although wound exudate plays a crucial role in the healing process by providing a moist wound environment, high exudate levels may delay ulcer healing, especially in hard-to-heal ulcers. Dressing types and their mode of action are critical factors that can influence clinical practice. There is a wide range of dressing technologies available to manage highly exuding LUs, such as alginates, hydrofibers, foams, and superabsorbents. Chronic wounds have elevated levels of metalloprotease (MMPs) activity, which are detrimental to healing (13). Studies have shown that superabsorbent dressings can bind and inhibit MMP activity, both in vitro (14, 15) and ex vivo (15), and that SAPs also have a high binding capacity for another proteinase, polymorphonuclear elastase (16). Some of the essential properties of SAPs include their ability to be effectively used under compression, providing cushioning, and, with a silicone layer/border, reducing the risk of damaging the skin and pain during dressing removal (17). Dressing selection should be consistent with wound-bed objectives and patients’ clinical needs. In addition, it has been shown that SAPs have a optimal benefit-harm (18) and cost-effectiveness profile in France (19), Germany (20), and UK (21). Superabsorbent wound dressings are endorsed for the management of moderate-to-high exudate chronic wounds by multiple clinical guidelines, position papers and consensus documents (17) (22). In addition to their use as secondary dressings for moderate-to-high exuding wounds, the WUWHS 2019 consensus document also suggest SAPs may be used as primary dressings for moderate-to-high exuding wounds (12).

The management of highly exuding wounds in patients requires a comprehensive approach as complications can arise throughout the healing process, resulting in significant impact on patient health and healthcare resource utilization (23). To allocate resources effectively, robust budget impact and cost-effectiveness analyses are necessary to aid the decision-making process (24). Therefore, it is crucial to assess the health economic impact of new wound dressing products, particularly given the increasing prevalence and economic burden of leg ulcers and the wide range of dressing options available. However, despite this pressing need, the number of full economic evaluations that compare different dressing technologies remains limited. Recent systematic literature reviews indicate that no studies have assessed the full economic impact of superabsorbent polymers (SAPs) for complex chronic wounds in the Spanish setting. Consequently, this study seeks to determine the cost-effectiveness/utility of using SAPs versus foam dressings in the management of moderate-to-highly exuding LUs and associated budget-impact analysis in Spanish settings, from the perspective of the Spanish National Health Service (NHS, composed of 17 regional health services as the 17 Spanish autonomous communities have full competences in health care).

## Methods

### Methodological Recommendations Standards

The economic evaluation adhered to both Spanish (25) and international guidelines (26), and the reporting quality was assessed based on the CHEERS checklist (27), along with the Drummond checklist to ensure adequate quality of conduct (28). Cost-effectiveness analysis was used as a vehicle for budget-impact analysis, which adhere to international (29, 30) and Spanish guidelines for budget impact analyses (31). Willingness-to-pay threshold (WTP) of 25,000 Euros was used in line with recent recommendations for Spain (32).

Systematic reviews were reported in accordance with the PRISMA statement (33), and the quality of evidence was appraised using the AMSTAR 2 critical appraisal tool (34). The evidence synthesis process involved the identification and quality appraisal of the most recent and highest level of evidence research. The identified data inputs were adjusted in line with recommendations for good research practices (35) and incorporated into a mathematical model that followed the natural history of chronic leg ulcers.

### Intervention and Comparator Dressings

The intervention employed for the evaluation was a superabsorbent dressing (Zetuvit Silicone Border, HARTMANN GROUP). As a mix of foams is commonly used for the treatment of moderate-to-highly exuding LUs, various types of foams were chosen as comparators to ensure a fair comparative assessment (a list of foams is provided in the supplementary file, Table S1).

The efficacy of SAP dressings was derived from individual patient data (IPD) obtained from two clinical trials evaluating the efficacy SAP dressings for the management of moderate-to-highly exuding leg ulcers (84 patients with a follow-up period of 2 weeks) (36, 37).

A systematic search was conducted in Embase and Medline vi ProQuest DIALOG, and Cochrane to identify studies reporting on the efficacy of foam dressings in reducing wound size after two weeks, as an outcome measure, in patients with moderate-to-highly exuding leg ulcers. This selection criterion was adopted to ensure a homogenous population with the subjects enrolled in clinical trials evaluating the efficacy of SAP. Our search retrieved 5,900 records, which were screened based on the title and abstract. Subsequently, 25 papers were identified for full-text screening. Ultimately, only three studies met the eligibility criteria and were included in the final analysis.

The present meta-analysis incorporated the aforementioned studies with the aim of quantifying the suitable decision-analytic model inputs for the efficacy of foams. The outcomes are depicted in the form of a forest plot, utilizing the mean difference as the metric for effect estimation. The statistical software R package “meta” was employed to conduct the meta-analysis. Given the continuous nature of the data (mean difference), the “metacont” function was implemented along with the integrated summary function “sm” and Hedges’ adjusted g-statistic was utilized.

In order to ascertain the efficacy of foam dressings with regards to wound size, calibration factors were calculated. These factors were derived from the research conducted by Szewczyk et al. (40) and are presented in tabular form below.

### Modelled Population Cohorts

The population characteristics used in this study were derived from individual patient data (IPD) obtained from two clinical trials evaluating the efficacy of superabsorbent dressings for the management of moderate-to-highly exuding leg ulcers (36, 37). The analysis included 84 patients with a follow-up period of 2 weeks (Table 1.). To ensure the stability of the results, 1000 patient profiles were generated per arm by random sampling from the original dataset. In the base-case analysis, multiple imputations of mean values were used to account for the small number of missing values in the baseline patient characteristics. For scenario analyses, missing values were inputted using various methods, including regression imputation and k-nearest-neighbor.

**Table 1.** Systematic review of foams dressing efficacy studies in the management of moderate-to-highly exuding leg ulcers

| Study | Age | SS | Gender | Wound duration | Wound size baseline | WAR (Day 14) | Absolute WAR/day | Relative WAR/day |
| --- | --- | --- | --- | --- | --- | --- | --- | --- |
| Vanscheidt et al. (38) | 66.40 | 46 | 50% | 10.91 | 1013 | 170 | 12.14 | 1.20% |
| Charles et al.(39) | 71.00 | 31 | 48.40% | 32 | 881 | 273.11 | 19.51 | 2.21% |
| Szewczyk et al.(40) | 62.20 | 20 | 40.00% | 17.5 | 1885 | 746.46 | 53.32 | 2.60% |

Legend: WSR – Wound Area Reduction, SS: sample size

For budget impact analysis we have used Berenguer et al. (5) latest results of 2.1 cases per 1000 persons per year and population 2023 census data from Spanish National Statistics Institute (Instituto Nacional de Estadísitica) of 47,615,034 people (41).

### Chronic Wound Model

The microsimulation model utilized in this study employs patient data obtained from clinical studies (36, 37) and allocates fundamental patient characteristics to each modelled patient as in the original primary data. The patients are generated with baseline characteristics (patient-related and wound-related). Those baseline patients and wound characteristics are the main predictors from Margolis et al. (42) risk prediction model. In the next step, the generated patient is cloned into two identical copies and then tested in the model to observer what will happen if they continue to be treated with SAP, or treated with foams from the start, with the only difference that SAP arm will have added two weeks of wound duration. In the next step, natural history was modelled for every patient and nested risk prediction model has predicted a unique patient trajectory for 24 weeks. At the end of the period (or earlier in case the patient having died), the accumulated Quality-Adjusted Life Weeks (QALWs) and healing status will be calculated. This process is executed to ensure that there is no disparity in patient or ulcer characteristics, and only the treatment assignments differ between the two arms. Margolis et al.(42) use logistic regression analysis for their risk prediction model. We calculate the risk in the following manner:

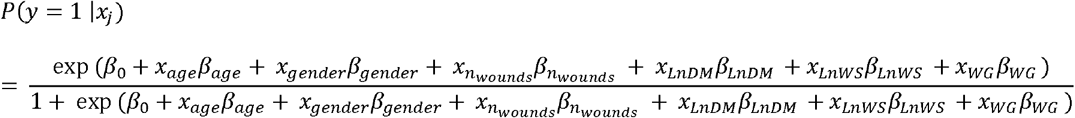

Legend: p denotes the probability, β0 the intercept, β the regression coefficient, x the independent variable, wounds – number of wounds, LnDM logarithm of wound duration in months, LnWS logarithm of wound size, and WG denotes the wound grade.

As shown in Figure 2, every patient from both arms progresses through various health states.

**Figure 1.**
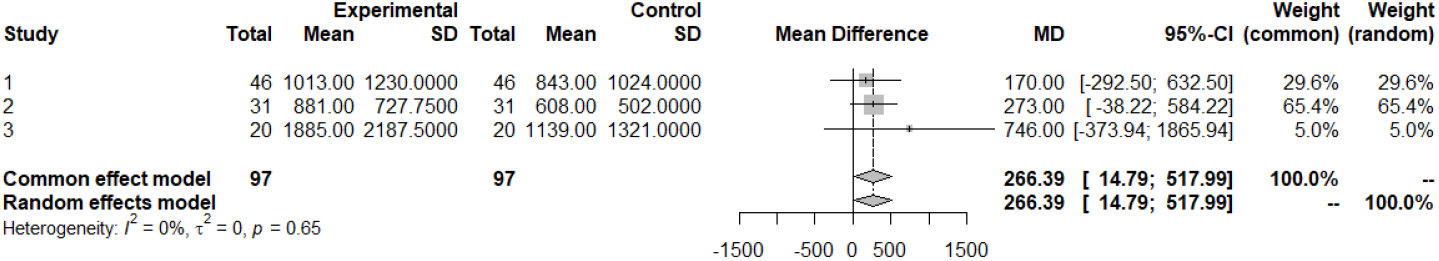
Foam dressings efficacy in reduction of wound size (mean difference baseline – 14 days follow-up).

**Figure 2.**
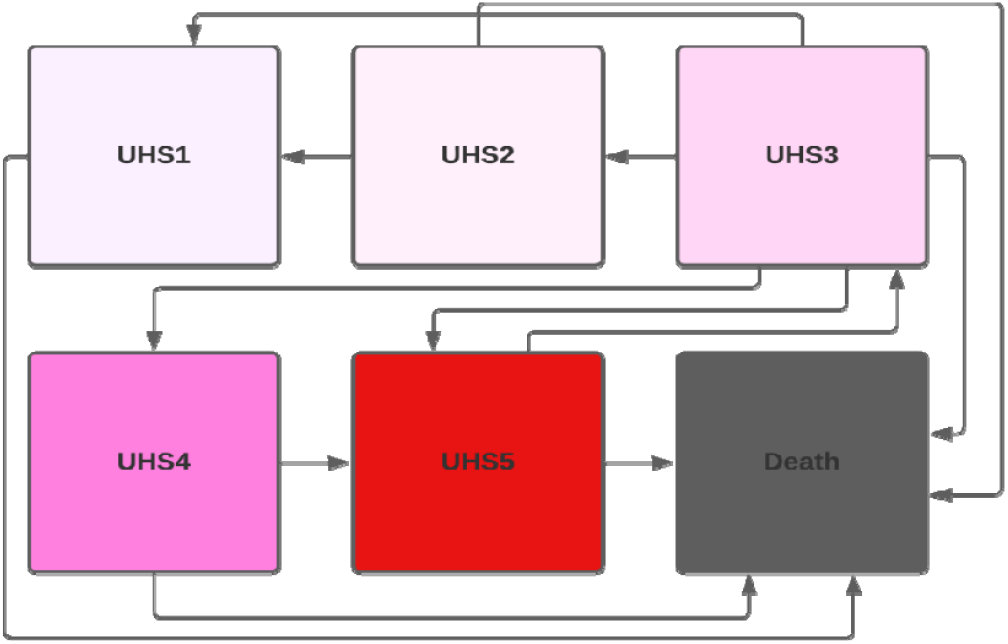
Decision analytic model diagram UHS1: Healed, UHS2: Unhealed grade 1: progressing, UHS3: Unhealed grade 1: static, UHS4: Unhealed grade 1: deteriorating, UHS5: Unhealed grade 2: severe, D: Death; *Adjusted for all patient using the risk prediction model

**Figure 3.**
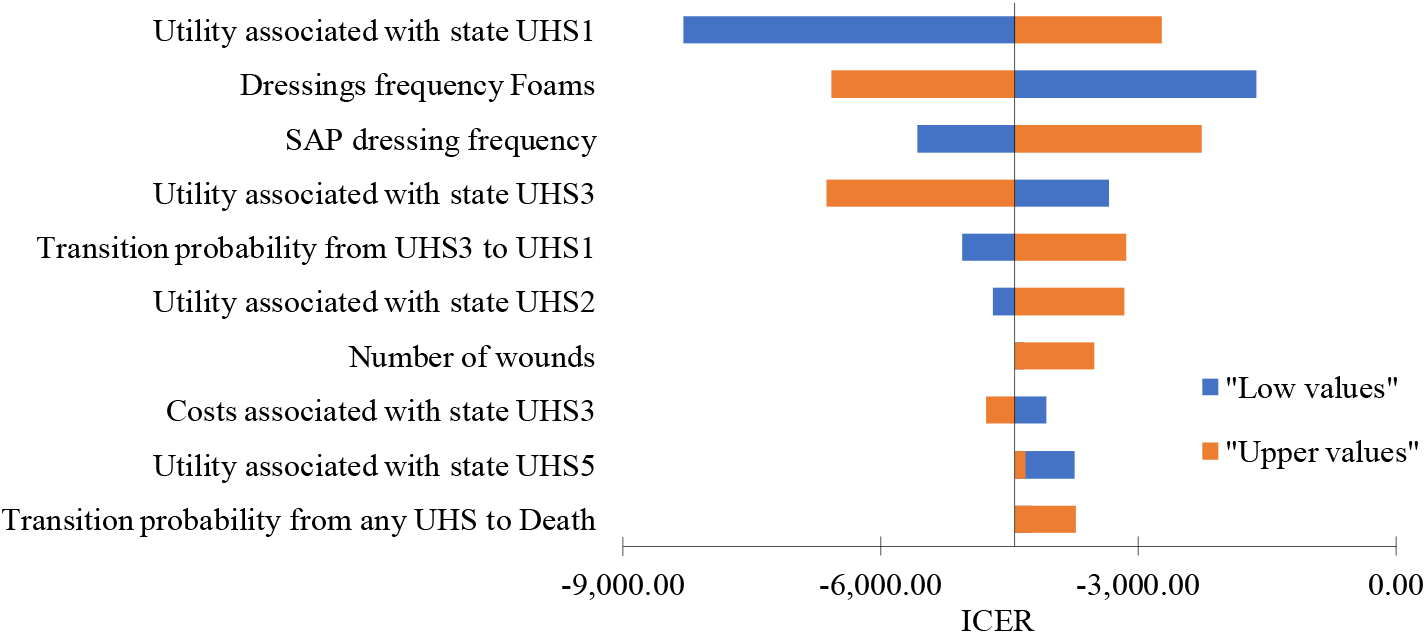
UHS1: Healed, UHS2: Unhealed grade 1: progressing, UHS3: Unhealed grade 1: static, UHS5: Unhealed grade 2: severe, SAP: Superabsorbent Wound Dressings, ICER: Incremental Cost-effectiveness ratio.

The final results are calculated as the average over the entire modelled population after 1000 generated patients have passed through the model for both treatment arms. Patients are classified into different ulcer health states (UHS), such as “healed: skin is intact” (UHS1), “Unhealed: grade 1 progressing: ulcer is progressing towards healing” (UHS2), “Unhealed grade 1 static: ulcer is neither healing nor deteriorating” (UHS3), “Unhealed grade 1 deteriorating: ulcer is deteriorating” (UHS4), and “Unhealed grade 2 severe: ulcer is infected or with other complications which may require hospital admission and/or surgical intervention” (UHS5). For transitions from “UHS3” to “UHS1” and transition probability from “UHS5” to “UHS3”, multiple sources from Norman et al. (43) were used for derivation, and in that case, we have applied following steps: (i) the probabilities are first identified and extracted, (ii) the probabilities of different lengths are transformed in the weekly probabilities, and (iii) weighted average was calculated for weekly probabilities using studies sample size as weight (details provided in the supplementary Tables S2/S3). From any health state, a patient may transition to the state of “Death”. The probabilities of transition from the remaining health states of the ulcer are uniform across time and arms (Table 2). The model’s weekly cycle was chosen to adequately reflect significant clinical changes in the progression of leg ulcers.

**Table 2.** Calibration factors per wound size category

| Wound size category | Calibration factor |
| --- | --- |
| Wound size category 1 (0-10 cm<sup>2</sup>) | 1.124 |
| Wound size category 2 (10-20 cm<sup>2</sup>) | 0.842 |
| Wound size category 3 (>21 cm<sup>2</sup>) | 1.035 |

Given that LUs are heterogeneous conditions, and the rate of healing depends on a wide range of factors, we considered the microsimulation model as the only relevant method to reflect all aspects of the pathology progression (26). The model flow and structure of the model health states replicate the natural history of the wound healing process (Figure 2).

### Cost Inputs

In this study, we employed the suggested cost methodology for chronic wounds to determine the cost per health state (47). Despite conducting a systematic review on cost-of-illness studies for Spain, we did not find any appropriate study, either with or without the suggested methodological approach. Consequently, we applied the corresponding Spanish cost to all resource use elements of chronic wound care, as recommended in the cost methodology paper by Harding et al. (47), to derive the baseline cost of static ulcer (UHS3). To estimate the appropriate cost for other health states, we derived calibration factors characterizing the difference in cost between different health states from the same method paper (Table 3) (breakdown of procedure and medicinal product costs in Spanish settings for the estimation of health state reported in the supplementary file, Table S4 and S5).

**Table 3.** Patient characteristics

| Variable | Mean (observations) | 95% CI |  |
| --- | --- | --- | --- |
| Age | 72.79 (78) | 69.88 | 75.69 |
| Gender (%) | 86.90 (83) | 71.55 | 100 |
| Number of wounds | 1 (84) | - | - |
| Duration of wounds (months) | 15.09 (78) | 8.34 | 21.84 |
| Wound size SoC (mm2): baseline | 5765 (84) | 662 | 10.868 |
| Wound size SoC (mm2): after two weeks | 3578 (84) | 1923 | 5.233 |

Legend: observations – number of observations, CI – confidence interval

The cost of dressings was considered separately and not included in the cost of the health states. The size of the dressings was matched with the wound size of each patient in the model to recreate similar clinical courses (product cost reported in the supplementary file, Table S6). The average cost per dressing by type was estimated from reference sources, and the number of dressing changes per type of dressing was informed by Panca et al. (44). We used Spanish-specific cost data inputs in Euros for all cost calculations in the model. We did not apply a discount rate for cost and outcomes as the time horizon was less than one year.

### Patient Outcomes

The health outcome measure recommended by the Spanish recommendations (25) is quality adjusted life years (QALYs). Therefore, in this study, we used a utility outcome measure called quality-adjusted life week (QALW), given the six-month time horizon. QALWs were computed by multiplying the utility value for each week gained (informed by age and gender-specific Spanish life tables) with the corresponding duration of one week, as measured in the primary studies (Table 4).

**Table 4.** State-transition model probabilities

| Transition probability | Value | Source |
| --- | --- | --- |
| Transition probability from “UHS2” to “UHS1” | 0.0250 | Panca et al. (44) |
| Transition probability from “UHS3” to “UHS2” | Patient specific | Risk-prediction model Margolis et al. (42) using IPD |
| Transition probability from “UHS3” to “UHS4” | 0.0188 | Walzer et al. (45) |
| Transition probability from “UHS3” to “UHS1” | 0.0188 | Meta-analysis based on Norman et al. (36) |
| Transition probability from “UHS4” to “UHS5” | 0.0040 | Shannon et al. (46) |
| Transition probability from “UHS3” to “UHS5” | 0.0170 | Walzer et al. (45) |
| Transition probability from “UHS5” to “UHS3” | 0.8000 | Meta-analysis based on Norman et al (43) |
| Transition probability from any health state to death | Age and gender-specific | Spanish life tables (41) |

Legend: UHS1: Healed, UHS2: Unhealed grade 1: progressing, UHS3: Unhealed grade 1: static, UHS4: Unhealed grade 1: deteriorating, UHS5: Unhealed grade 2: severe, D: Death; IPD: Individual Patient Data, \*Adjusted for all patient using the risk prediction model

The utility values ranged from zero (representing death) to one (representing perfect health) and were informed by evidence from published literature (48, 49). Furthermore, we assessed the utility values in accordance with international practice recommendations (50), which are presented in the table above. The primary effectiveness outcome measure chosen for this study was the healing rate. We evaluated cost-effectiveness by identifying the interventions on the cost-effectiveness frontier and presenting the results using the incremental cost-effectiveness ratio (ICER) and the net monetary benefit (NMB) to facilitate interpretation.

### Time Horizon and Perspective

In accordance with the observed consequences and relevant outcomes of the dressing therapy, a time horizon of six months (24 weeks) was deemed appropriate for capturing differences in costs and health effects. A lifetime horizon was deemed inadequate as wound dressings are unable to influence patient survival or modify the natural history of the underlying condition in any significant manner. Furthermore, currently available data do not permit acceptable extrapolation to a lifetime horizon or any timeframe longer than six months. The economic evaluation was conducted from the perspective of the third-party payer, the NHS, and therefore only healthcare system resources involved in the production of patient care and health effects were taken into consideration (25). There was not sufficient publicly available data for Spain to conduct the analysis using broader societal perspective.

### Uncertainty Quantification

To evaluate the uncertainty associated with the structural choices of the evaluation and their impact on the results, we conducted two types of sensitivity analyses: one-way sensitivity analysis (OWSA) and multivariate probabilistic sensitivity analysis (PSA) (51).

In OWSA, we set each parameter at its extreme upper and lower bounds, while the rest of the parameters remained fixed, to assess the impact on the results. For parameters without available confidence interval (CI) values, we used ±20% point estimate for the lower and upper bounds. PSA enabled us to assess the joint effect of uncertainty over all parameters. We varied all parameters simultaneously according to their plausible values by random sampling. We repeated the process of sampling inputs and recording outputs 5000 times to obtain a reliable estimate of expected costs and effects. Given that PSA employs parameters as distributions of possible mean values instead of single point estimates, we selected the distribution in line with the recommendations of good modelling practice (35). We used the method of moments to estimate parameters for the distribution. The distributions used were normal, beta, gamma, and Dirichlet distribution (Table below). Selection of distributions per parameters are based on the existing recommendations 2. Parameters for distributions were constructed using methods of moments. For inputs lacking the precise inputs in terms of variability measures, we used the standard deviation of 0.5.

## Results

According to the findings of our early-stage health economic evaluation, substitution of foam dressings with SAP in patients with moderate-to-highly exuding leg ulcers in a Spanish setting would result in an enhanced healing rate of 2.33%, incremental HRQoL of 0.129 QALWs, and total direct cost savings of € 570 per patient over a period of six months. In other words, using SAP dressings as an adjunct to compression therapy would result in an additional improvement of 2.33% in wound healing rate, leading to cost savings of € 570 per patient. Additionally, patients would experience an incremental quality of life equivalent to one extra day of perfect health over the course of six months. At the end of the six-month period, 33.25% of patients treated with SAP would achieve wound closure, compared to 30.92% of patients treated with foams dressing mix.

Total national level budgetary impact for the same period is 54,11 million of Euros. In other words, according to findings of this analysis in scenario in which all LUs will be treated with SAPs instead of foams, Spain NHS can have annual saving of 43,46 million of Euros (Table below).

To examine the impact of input variable uncertainty on the results, we conducted a one-way deterministic sensitivity analysis (OWSA) by testing all variables included in the model. The OWSA was performed by running the model on both the lower and upper ranges/confidence intervals of each parameter while holding all other parameters constant. The procedure was repeated for each variable in the model, and the results were presented in tornado diagram, which depicted the ten most influential parameters on incremental cost-effectiveness ratio (ICER). The OWSA demonstrated that the deterministic results were robust, and the conclusions from Table 5 remained unchanged.

**Table 5.** Cost per health state

| Health state | UHS1 | UHS2 | UHS3 | UHS4 | UHS5 |
| --- | --- | --- | --- | --- | --- |
| Cost per health state | 7.69 € | 111.54 € | 128.21 € | 159.45 | 637.15 |

Legend: UHS1: Healed, UHS2: Unhealed grade 1: progressing, UHS3: Unhealed grade 1: static, UHS4: Unhealed grade 1: deteriorating, UHS5: Unhealed grade 2: severe, D: Death; \*Adjusted for all patient using the risk prediction model

**Table 6.** Utility values per health state

| Utilities per health state | Value | Source |
| --- | --- | --- |
| UHS1 | 1.000 | Clegg et al. (48) |
| UHS2 | 0.730 |  |
| UHS3 | 0.640 |  |
| UHS4 | 0.640 |  |
| UHS5 | 0.610 | Assumption based on Matza et al. (49) |
| D | 0 | NA |

Legend: UHS1: Healed, UHS2: Unhealed grade 1: progressing, UHS3: Unhealed grade 1: static, UHS4: Unhealed grade 1: deteriorating, UHS5: Unhealed grade 2: severe, D: Death, NA – non applicable.

**Table 7.**
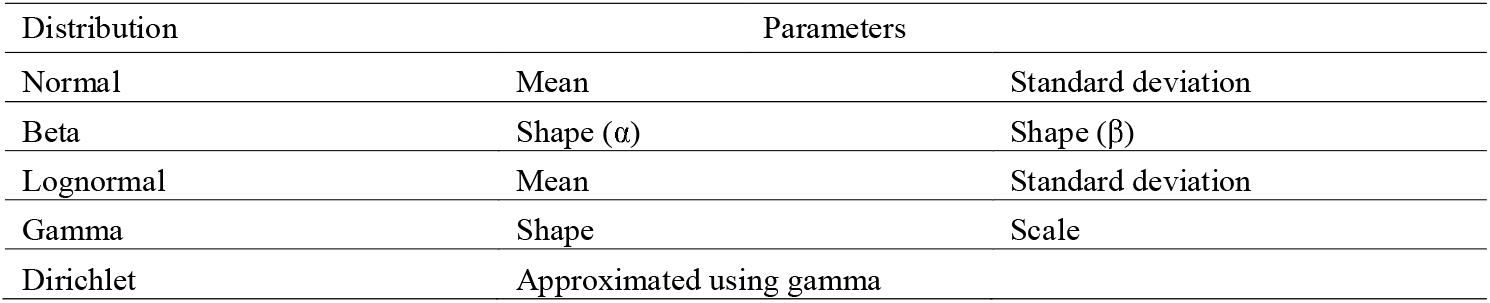
Parameters for distributions

**Table 8.** Results of cost-effectiveness analysis

| SAPs |  |  | Foams |  |  | Incremental Costs | Incremental QALWs | Incremental HR | ICER (QALY) | NMB |
| --- | --- | --- | --- | --- | --- | --- | --- | --- | --- | --- |
| Cost | QALWs | HR | Cost | QALWs | HR |  |  |  |  |  |
| 3,836 € | 17.161 | 33.25% | 4,407 € | 17.032 | 30.92% | -570 € | 0.129 | 2.33% | -4,406. € | 3,807 € |

Legend: SAPs: Superabsorbent Wound Dressings, QALWs: Quality Adjusted Life Weeks, HR: Healing Rate, ICER: Incremental Cost-effectiveness ratio, NMB: Net Monetary Benefit (under WTP threshold of 25,000 Euros).

**Table 9.** Results of cost-effectiveness analysis

| Prevalence | Census Spain, N | Patients with wounds, N | Moderate-to-highly Exuding Leg Ulcers, N | Total Cost of treating all patients with |  | Budget Impact |
| --- | --- | --- | --- | --- | --- | --- |
|  |  |  |  | SAPs Dressing | Foams Dressing |  |
| 0.8% | 47,615,034 | 380,920.27 | 76,184.054 | 292,267,802 € | 335,723,527 € | -43,455,725 € |

Legend: N – total population, SAPs: Superabsorbent Wound Dressings

We also conducted a PSA, which demonstrated that the use of SAP dressings was a cost-saving option and resulted in higher benefits in 100% of cases. We provided detailed results of the probabilistic analysis in the supplementary material (Table S5 and Figure S1).

## Discussion

To the best of our knowledge, this study is the first attempt to estimate the cost-effectiveness of superabsorbent polymer (SAP) wound dressings in the Spanish Healthcare System scenario, and the first model-based evaluation of wound dressings in chronic wounds. Our study conducted a detailed exploration of five published systematic literature reviews of economic evaluations and cost-of-illness studies, aiming to identify economic evaluations in Spanish settings. However, those systematic literature reviews did not identify any economic study for chronic ulcers in Spanish settings.

This study lays the groundwork for further investigations in a critical area with a conspicuous research gap. To this end, we conducted a comparison of our findings with the most comparable studies in France, United Kingdom and Germany, utilizing the same time horizons and cost of SAPs and control arm inflated for the same year (2021) and currency. Our results appear to be in line with the average results of other economic evaluations. This is unsurprising since our study adheres to international recommendations for data selection and economic evaluations, follows the recommended methodology for costing chronic leg ulcers, and builds on the latest published models.

Despite following recommended research practices, there are limitations to this study that must be acknowledged to properly understand its implications for decision making. Clinical management guidelines advocate for the use of superabsorbent dressings (SAPs) as the primary choice for highly exuding ulcers, despite the lack of confirmation of efficacy through robust randomized controlled trials (RCTs). While our comparison was done in a counterfactual manner using the same population as a basis, the estimation of treatment effects was based on non-randomized trials. Consequently, unmeasured confounding could not be adjusted at the mathematical/biostatistical analysis level, and the treatment effect may be biased by an unknown amount. To address this shortcoming to some extent, we conducted several deterministic and probabilistic scenario analyses to test the robustness of the results. According to the results of these sensitivity analyses, the conclusions based on the base-case analysis remained unchanged in every plausible range and every combination of input parameters used. However, further RCTs are needed to confirm these findings definitively. It should be noted that we used the most conservative scenario for SAPs, and future RCTs are expected to demonstrate even more significant clinical effects of SAPs compared to foams in this population. Thus, the current model should be considered as an early health technology assessment of SAPs, owing to the immature non-randomized clinical efficacy data that is used as input parameters for this model, which has the potential benefit. In situations where the lack of RCT data results in a high degree of uncertainty in medical decision-making, decision-analytic modelling is recommended to provide decision support to clinical practitioners and payors.

## Conclusion

According to the findings of this preliminary health economic analysis, the use of SAP wound dressings in comparison with foam dressings mix in patients with moderate-to-highly exuding leg ulcers in Spanish Health Care System scenario is predicted to result in improved healing rate of 2.33%, an incremental HRQoL of 0.129 QALWs, and total direct cost savings of € 570 per patient over a six-month period. According to this analysis’s findings, in a scenario in which all LUs will be treated with SAPs instead of foams dressings, Spain’s NHS can have an annual saving of 43,46 million Euros.

These clinical outcomes align with current management guidelines that recommend superabsorbent wound dressings as a primary option for the therapy of moderate-to-highly exuding leg ulcers. Furthermore, the evaluation demonstrates that the use of superabsorbent wound dressings can also lead to economic savings from the perspective of the NHS in Spain.

## Data Availability

All avaiable data are reported.

