## Supplementary material for "Management of Moderate-to-highly Exuding Leg Ulcers with Superabsorbent Wound Dressings versus Foams Dressings in Spanish Settings: An Early-stage Cost-effectiveness and Budget-impact Analyses": Suplementary File: Supplementary file_Final.pdf

Table S1. Mix of foams dressings in the comparator arm

| Foams | Size (N) | Manufacturer |
|---|---|---|
| Mepilex Border Flex 15x15 | 5 | Mölnlycke |
| Mepilex Border Flex 7,5 x7,5 | 5 | Mölnlycke |
| Mepilex Border Flex 10x10 | 5 | Mölnlycke |
| Mepilex Border Flex 20x20 | 5 | Mölnlycke |
| Mepilex 10x10 | 5 | Mölnlycke |
| Mepilex 15x15 | 5 | Mölnlycke |
| Allevyn Gentl border 7,5x 7,5 | 10 | Smith & Nephew |
| Allevyn Gentler border 12,5x12,5 | 10 | Smith & Nephew |
| Allevyn Gentler border 10x20 | 10 | Smith & Nephew |
| Biatain Silicone 7,5x7,5 | 10 | Coloplast |
| Biatain Silicone 10x10 | 10 | Coloplast |
| Biatain Silicone 12,5x12,5 | 10 | Coloplast |
| Biatain silicone 15x15 | 5 | Coloplast |
| Biatain Silicone 17,5x17,5 | 5 | Coloplast |
| Allevyn Life 10,3 x10,3 | 10 | Smith & Nephew |
| Alevyn life 15,4x 15,4 | 10 | Smith & Nephew |
| Allevyn Life 21x21 | 10 | Smith & Nephew |

Table S2. Transition probability from HS3 to HS1

| Study | Country | Size | Follow-up | Healing % | Weeks | Rate | Risk |
|---|---|---|---|---|---|---|---|
| Fogh, 2012 | Europe | 60 | 1.5 months | 0.1833 | 6.51786 | 0.03106594 | 0.03058835 |
| Gottrup, 2008 | Europe | 60 | 1.5 months | 0.16 | 6.51786 | 0.0267501 | 0.02639548 |
| Harding, 2001 | UK | 65 | 12 weeks | 0.2615 | 12 | 0.02526118 | 0.02494479 |
| Norkus, 2005 | Denmark | 48 | 12 months | 0.4 | 52.14288 | 0.00979665 | 0.00974882 |
| Schulze, 2001 | Germany | 113 | 4 weeks | 0.05 | 4 | 0.01282332 | 0.01274146 |
| Meaume, 2012 | France | 94 | 8 weeks | 0.0745 | 8 | 0.00967764 | 0.00963097 |
| Senet, 2014 | Europe | 94 | 6 weeks | 0.0319 | 6 | 0.00540332 | 0.00538874 |
| Sopata, 2016 | Poland | 50 | 40 weeks | 0.76 | 40 | 0.03567791 | 0.03504895 |
| Vanscheidt, 2012 | Germany | 66 | 12 weeks | 0.32 | 12 | 0.03213854 | 0.03162759 |
| Weighted Average | | | | | | | **0.0187691** |

Table S3. Transition probability from HS3 to HS5

| Study | Country | Size | Follow-up | Healing % | Weeks | Infection | Rate | Risk |
|---|---|---|---|---|---|---|---|---|
| Fogh, 2012 | Europe | 60 | 1.5 months | 0.1833 | 6.51786 | 0.03 | 0.00467319 | 0.00466229 |
| Gottrup, 2008 | Europe | 60 | 1.5 months | 0.16 | 6.51786 | 0.03 | 0.00467319 | 0.00466229 |
| Harding, 2001 | UK | 65 | 12 weeks | 0.2615 | 12 | 0.0833 | 0.00724792 | 0.00722172 |
| Schulze, 2001 | Germany | 113 | 4 weeks | 0.05 | 4 | 0.2 | 0.05578589 | 0.05425839 |
| Meaume, 2012 | France | 94 | 8 weeks | 0.0745 | 8 | 0.0638 | 0.00824077 | 0.00820691 |
| Sopata, 2016 | Poland | 50 | 40 weeks | 0.76 | 40 | 0.12 | 0.00319583 | 0.00319073 |
| Vanscheidt, 2012 | Germany | 66 | 12 weeks | 0.32 | 12 | 0.091 | 0.00795085 | 0.00791932 |
| Weighted Average | | | | | | | | **0.016956** |

Table S4. Breakdown of procedure costs in Spanish settings for the estimation of health state costs

| Category | Type | DRG code | Fee per procedure |
|---|---|---|---|
| Investigations (outpatient) | Ultrasonography | III.3.1.2.1.5.1. | 36.92 € |
| | Angiography | III.3.1.2.2.7.1 | 443.04 € |
| | Phlebography | III.3.1.2.2.2.2 | 166.14 € |
| | Biopsy | III.3.1.1.3.14 | 27.51 € |
| | Allergy tests | III.3.4.21 | 48.69 € |
| **Category** | **Type** | **DRG code** | **Fee per procedure** |
| Outpatient clinic visits (fee per consultation) | Physiotherapist | III.8.3 | 23.64 € |
| | Therapist of lymphatology | III.8.2 | 52.86 € |
| | Nurse | I.3.1.1 | 24.69 € |
| | Practitioner | I.1.1.1 | 53.75 € |
| | Consultant | I.1.1.1 | 53.75 € |
| | Specialised outpatient clinic | I.1.1.1 | 53.75 € |
| **Category** | **Type** | **DRG code** | **Fee per procedure** |
| Hospital admission | Arterial surgery | III.1.1.130 | 4,748.33 € |
| | Vein Surgery | III.1.1.119 | 2,853.63 € |
| | Meshgraft transplantation | III.1.1.264 | 7,057.09 € |
| | Hospitalisation | III.1.1.271 | 6,008.52 € |
| | Conservative therapy and VAC | III.1.1.270 | 3,517.00 € |

Source of Costs: Ministerio de Sanidad, Consumo y Bienestar Social (Portal estadístico) / Ministry of Health, Consumption and Social Welfare (Statistical Portal)

Table S5. Breakdown of medical products costs in Spanish settings for the estimation of health state costs

| Category | Type | Typicaly used product | Price per pack |
|---|---|---|---|
| Skin care | Topical ointments | Hyperogenated fatty acids lotion | 8 € |
| | Skin protection | Crenas hidrotantes | 12 € |
| | Antibiotic ointments | Metrodinazol | 6 € |
| | Topical anesthetic ointment | Lidocaine (one vial, 2% and 10 ml.) | 0,33 € |
| Analgesics and antibiotics for wound | Anticoagulant | Mupirocina | 8 € |
| | Immunosupressant | Lidocaina Emla | 6 € |
| | Antibiotic | Amoxicillin with clavulanic acid (one vial i.v, 1 gr.) | 0,90 € |
| | Analgesic | Cortisona | 6 € |
| | Psycotropic | Trankimazin | 8 € |
| Compression hosiery | Compression stockings/Lower leg | 1 | 25 € |
| | Compression stockings/Upper leg | 2 | 45 € |
| | Compression stockings/Pelvis | 3 | 75 € |

Source of Costs: Grupo HLA (Departamento de Farmacia) / HLA Healthcare Group, Pharmacy Department (private group)

Table S6. Product costs

| SAPs: | Size (N) | Manufacturer | Price per package | Price per Dressing |
|---|---|---|---|---|
| Zetuvit Plus Silicone 12,5x12,5cm P10 | 10 | Hartmann | 23.63 | 2.788 |
| Zetuvit Plus Silicone 10x20cm P10 | 10 | Hartmann | 24.54 | 2.896 |
| Zetuvit Plus Silicone 20x20cm P10 | 10 | Hartmann | 45.45 | 5.363 |
| Zetuvit Plus Silicone Border 10x10cm P10 | 10 | Hartmann | 26 | 3.068 |
| Zetuvit Plus Silicone Border 12,5x12,5cm P10 | 10 | Hartmann | 27.9 | 3.292 |
| Zetuvit Plus Silicone Border 17,5x17,5cm P10 | 10 | Hartmann | 42.9 | 5.062 |
| Zetuvit Plus Silicone 12,5x12,5cm P10 | 10 | Hartmann | 23.63 | 2.788 |
| **Foams:** | **Size (N)** | **Manufacturer** | **Price per package** | **Price per Dressing** |
| Mepilex Border Flex 15x15 | 5 | Mölnlycke | 25.50 € | 6.018 |
| Mepilex Border Flex 7,5 x7,5 | 5 | Mölnlycke | 7.00 € | 1.652 |
| Mepilex Border Flex 10x10 | 5 | Mölnlycke | 14.00 € | 3.304 |
| Mepilex Border Flex 20x20 | 5 | Mölnlycke | 27.00 € | 6.372 |
| Mepilex 10x10 | 5 | Mölnlycke | 10.50 € | 2.478 |
| Mepilex 15x15 | 5 | Mölnlycke | 23.50 € | 5.546 |
| Allevyn Gentl border 7,5x 7,5 | 10 | Smith & Nephew | 0.80 € | 0.094 |
| Allevyn Gentler border 12,5x12,5 | 10 | Smith & Nephew | 1.40 € | 0.165 |
| Allevyn Gentler border 10x20 | 10 | Smith & Nephew | 3.80 € | 0.448 |
| Biatain Silicone 7,5x7,5 | 10 | Coloplast | 18.00 € | 2.124 |
| Biatain Silicone 10x10 | 10 | Coloplast | 19.00 € | 2.242 |
| Biatain Silicone 12,5x12,5 | 10 | Coloplast | 23.40 € | 2.761 |
| Biatain silicone 15x15 | 5 | Coloplast | 15.90 € | 3.752 |
| Biatain Silicone 17,5x17,5 | 5 | Coloplast | 16.50 € | 3.894 |
| Allevyn Life 10,3 x10,3 | 10 | Smith & Nephew | 29.00 € | 3.422 |
| Alevyn life 15,4x 15,4 | 10 | Smith & Nephew | 41.80 € | 4.932 |
| Allevyn Life 21x21 | 10 | Smith & Nephew | 67.00 € | 7.906 |

Source of Costs: Telecita tools

Table S7. Results of the PSA analysis

| | SAP | | Foams | | Incremental Costs | Incremental QALWs | ICER |
|---|---|---|---|---|---|---|---|
| | Cost± | QALYs | Cost | QALWs | | | |
| **Means** | 3,804 € | 17.161 | 4,379 € | 17.019 | -575 € | 0.142 | -4,064 € |
| **Medians** | 3,804 € | 17.164 | 4,401 € | 17.014 | -598 € | 0.150 | -3,973 € |

Figure S1. Cost-effectiveness plane

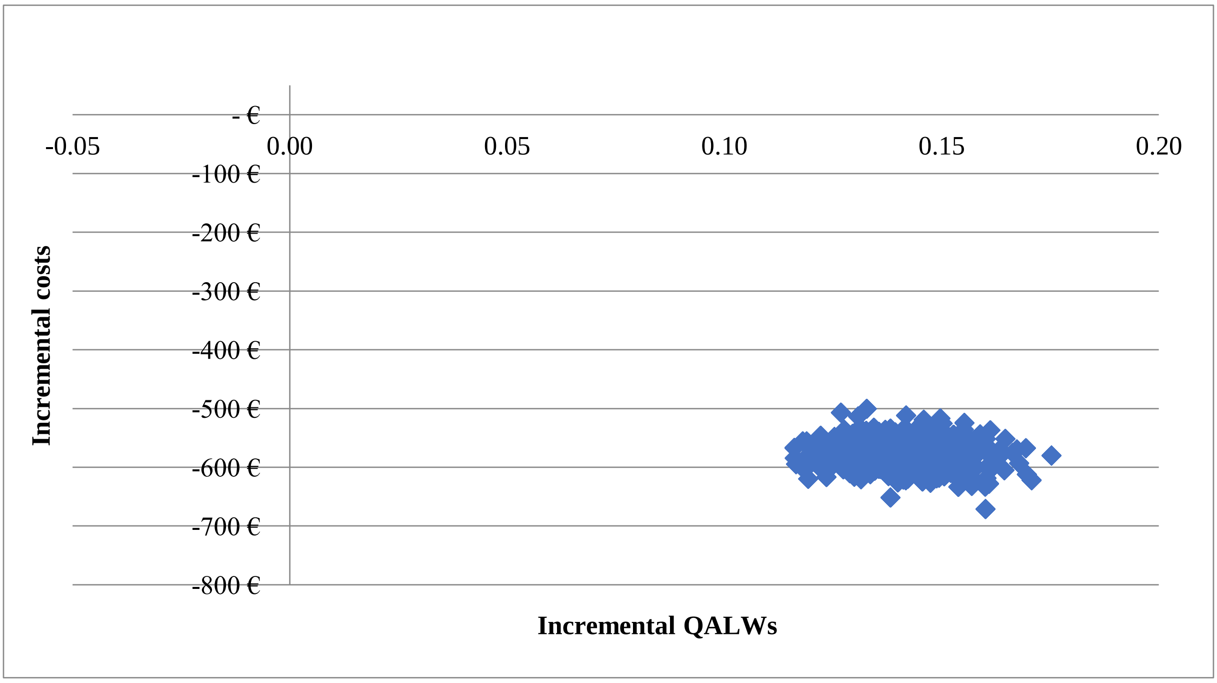
